# An mHealth app using machine learning to increase physical activity in diabetes and depression: clinical trial protocol for the DIAMANTE Study

**DOI:** 10.1101/2020.06.29.20142943

**Authors:** Adrian Aguilera, Caroline A. Figueroa, Rosa Hernandez-Ramos, Urmimala Sarkar, Anupama G Cemballi, Laura Gomez-Pathak, Jose Miramontes, Elad Yom Tov, Bibhas Chakraborty, Xiaoxi Yan, Jing Xu, Arghavan Modiri, Jai Aggarwal, Joseph Jay Williams, Courtney R. Lyles

## Abstract

**Introduction:** Depression and diabetes are highly disabling diseases with a high prevalence and high rate of comorbidity, particularly in low-income ethnic minority patients. Though comorbidity increases the risk of adverse outcomes and mortality, most clinical interventions target these diseases separately. Increasing physical activity might be effective to simultaneously lower depressive symptoms and improve glycemic control. Self-management apps are a cost-effective, scalable and easy access treatment to increase physical activity. However, cutting-edge technological applications often do not reach vulnerable populations and are not tailored to an individual’s behavior and characteristics. Tailoring of interventions using machine learning methods likely increases the effectiveness of the intervention.

**Methods and analysis:** In a three-arm randomized controlled trial we will examine the effect of a text-messaging smartphone application to encourage physical activity in low-income ethnic minority patients with comorbid diabetes and depression. The adaptive intervention group receives messages chosen from different messaging banks by a reinforcement learning algorithm. The uniform random intervention group receives the same messages, but chosen from the messaging banks with equal probabilities. The control group receives a weekly mood message. We aim to recruit 276 adults from primary care clinics aged 18 to 75 years who have been diagnosed with current diabetes and show elevated depressive symptoms (PHQ-8 >5). We will compare passively collected daily step counts, self-report PHQ-8 and most recent HbA1c from medical records at baseline and at intervention completion at 6-month follow-up.

**Ethics and dissemination:** The Institutional Review Board at the University of California San Francisco approved this study (IRB: 17-22608). We plan to submit manuscripts describing our User Designed Methods and testing of the adaptive learning algorithm and will submit the results of the trial for publication in peer-reviewed journals and presentations at (inter)-national scientific meetings.

**Registration:** clinicaltrials.gov: NCT03490253; pre-results

**Article Summary:** *Strengths and Limitations:*

- Novel approach of targeting diabetes and depressive symptoms using a smartphone application
- Ability to compare adaptive messaging for increasing physical activity to messages delivered with equal probabilities
- Testing of a smartphone application integrated within primary care settings in a low-income vulnerable patient population
- Longitudinal design with 6-month follow-up enables assessing intervention effects over time
- Challenges of this trial include supporting users in key behavior change in an automated manner with minimal in-person support

## INTRODUCTION

### Background

Both depression and diabetes are highly prevalent, often co-occurring diseases that are among the major causes of global disability (1,2,3). Comorbid diabetes and depression is associated with a worse prognosis of both diseases, including a higher rate of complications of diabetes, greater disability, and an increased risk of mortality (4). Vulnerable populations, including low-income, low health literacy and ethnic minority individuals experience higher prevalences and worse outcomes for both diabetes and depression.(5,6) There is a great need for the development of treatments that can target overlapping risk factors for diabetes and depression. A growing body of evidence suggests that physical activity is such a risk factor: it is linked to both mental health and diabetic outcomes (7,8,9,10,11)

Mobile applications have been found effective in helping patients engage in healthy behaviors including physical activity. For instance, a recent meta-analysis of nine RCT’s concluded that smartphone apps that focus on physical activity have a moderate positive effect on increasing physical activity levels, and another meta-analysis including 18 studies moderate to large effect in daily step changes(12,13). These effect sizes are similar to ‘face-to-face’ interventions(14). However, because around 70% of lower income Americans (including Latinos) currently own smartphones(15), and the ownership of smartphones is expected to increase in low income populations globally(16), mobile apps have great potential to reach individuals that normally do not have access to care. Further, mobile technology can help overcome existing barriers in access to care for vulnerable populations, including lower availability of psychological treatment in primary care settings, language and literacy barriers, stigma, cost, and inflexible employment schedules(17). Latinos in the US in particular show higher under-utilization rates of mental health treatment than non-Latino whites(18). Deploying effective mobile applications can therefore decrease existing decrease disparities in health.

However, in the U.S., low-income minority patients frequently receive their care in safety net settings (services in the public sector for those unable to attain private health insurance), where novel mobile technologies are not often designed, developed or implemented(19). This translational gap increases the probability that these interventions will ultimately fail when implemented in actual clinical settings that serve vulnerable populations.

Further, most mobile applications that target behavioral changes are not personalized(20), which could contribute to lower effect sizes of these interventions for trials with longer study durations (e.g. over 3 months)(12). Smartphone interventions allow for data collection by passive sensing technologies, which offer an opportunity for tailoring and personalizing interventions to users’ behaviors, preferences and needs. Personalization can be achieved by computer tailoring: tailoring interventions to observed behavior and characteristics of the participant. This can include feedback, goal setting, or user targeting (i.e. conveying that communication is designed specifically for the user)(21). However, more complex forms of personalization might be needed to increase and maintain engagement with PA interventions (22), a requirement for a digital intervention to be effective.

One promising approach is to use adaptive learning, which allows the prediction of which content might be effective for users, learning from its previous actions and participant data collected by mobile phones(23). For instance, an adaptive learning algorithm might be more effective as it can match motivation needs for each participant. Some intervention studies have attempted cultural tailoring to large groups of people. However, most tailoring approaches occur at the group level whereas we are bringing the tailoring down to the individual level(24). Research on the efficacy of these adaptive treatments is still in its early stages.

### Aim and hypotheses

The main aim of the “Diabetes and Mental Health Adaptive Notification Tracking and Evaluation” trial (DIAMANTE) is to test a smartphone intervention that generates adaptive messaging, learning from daily patient data to personalize the timing and type of text-messages. We will compare the adaptive content to **1**. a uniform random messaging intervention, in which the messaging content and timing will be delivered with equal probabilities (i.e. not adapted by a learning algorithm) **2**. a control condition that only delivers a weekly mood message. The primary outcomes for this aim will be improvements in physical activity at 6-month follow-up defined by daily step counts.

We will investigate the following specific hypotheses:

#### Primary Hypothesis

We expect that the group receiving adaptive messaging will have a statistically significant higher increase in step counts over the 6-month intervention period, compared to both the group receiving the uniform random messaging and the control group receiving only a weekly mood message. We expect that both the adaptive and the uniform random messaging groups will have a higher increase in step counts than the control group.

#### Secondary hypotheses

- We expect that the group receiving adaptive, personalized messages will have significant reduction in HbA1c levels, depression scores, weekly mood ratings, higher behavioral activation, lower rumination and anxiety, more positive attitudes about physical activity and higher self-efficacy to manage chronic diseases at 6 months, compared to the group that receives the uniform random messaging intervention and the control group.
- Exploratory: Because of the unique nature of the patient population enrolled in our study, we expect to find differences by language (English vs. Spanish). Language has been shown to be especially influential in mobile technology uptake and effectiveness in our previous work(25). Spanish speakers may engage more in the mobile intervention and might have different motivations for becoming physically active, which might result in differences in effectiveness of the various messages.

## METHODS AND ANALYSIS

### Design

This study is a randomized, controlled, single-center superiority trial with three groups and a primary endpoint of increase in daily number of steps during a 6-month intervention delivered by a smartphone app. Randomization will be performed as block randomization with a 1:1:1 allocation. Patients will be automatically randomized into groups through our secure server during onboarding of the app, thereby ensuring allocation concealment. Patients will be informed of the nature and frequency of the messages they will be receiving and will be allowed to discuss this with investigators during the course of the study.

Further, if messages are not being sent out appropriately, research assistants will contact the app developer to address errors within 24 hours. If physical activity data is not coming in, they may need to contact participants to ensure that the app is actively running on their phone, and assist participants with re-downloading the app if necessary. The necessity of these steps makes it unfeasible to blind the researchers. We used the SPIRIT checklist when writing our report(26).

### Population

Patients will be recruited within various clinics in the San Francisco Health Network, including the Richard Fine People’s Clinic, the Family Health Center, and from several endocrinologist providers at the Zuckerberg San Francisco General hospital (ZSFG); as well as the Department of Public Health in San Francisco, California in the United States that will serve as the study sites for this trial. These clinics serve a diverse, chronically ill population and, as in many public hospital settings, they employ multiple part-time providers, creating the risk for lack of continuity of care that mobile health programs may be able to supplement.

#### Inclusion criteria

Patients aged 18 to 75 years who have been diagnosed with diabetes and documented depressive symptoms (score > 5 on the PHQ-8), as determined by their medical health records, will be eligible for this study. If the patients’ health records show a diagnosis of diabetes but do not include information on PHQ-8 scores or depression diagnosis, we will assess patients’ PHQ-8 scores by telephone to determine their eligibility. After the assessment, this score will be uploaded into the patient health records, regardless of their eligibility for the study. Further, the researchers will reach out to the patient and his/her Primary Care Physician if participants screen > 20 on the PHQ-8 (severe depression). Eligible patients will need to use text messaging and have an iPhone or Android smartphone in order to download the pedometer app onto their phones, but they do not need to be proficient in using mobile phone applications. Concomitant care and interventions are permitted during the trial.

#### Exclusion criteria

We will exclude patients with: an inability to exercise due to physical disability; active psychosis or mania; active suicidal ideation; severe cognitive impairment; inability to read and write in English or Spanish; current pregnancy; plans to leave the country for extended periods of time during the 6-month trial.

#### Recruitment

Patients will be recruited using flyers placed in the waiting rooms, direct provider referrals and in-person recruitment strategies timed to visits. To identify potentially eligible patients, we will ask for permission from known providers to pull patient lists, and review and identify patients with dual diagnosis of diabetes and depression that would fit our inclusion criteria (e.g. is able to walk and is not pregnant).

#### Baseline visit

A researcher or healthcare provider will ask patients if they are interested in joining a study to increase physical activity and help manage their mood using their mobile phone. We will bring interested individuals into our offices at ZSFG for a baseline study visit and to obtain informed consent in Spanish or English. Thereafter we will collect all baseline survey measures of interests (outlined below) as well as patient demographics and information about current mobile technology familiarity and utilization. All patients will receive assistance in downloading a pedometer application onto their phone and will send test text-messages back to our system. The researcher will construct a plan for physical activity goals with the patient and instruct patients to have the app open at all times. Thereafter, users will be automatically randomized by the secure server, using a block randomization with block size 3 to allocate individuals into either the uniform random, adaptive, or control group.

#### Partial Patient and Public Involvement

Patients worked with us to design the text-messages, mobile app user interface, and were asked to assess the burden and time-commitment of the study, as part of our User Centered Design approach. We did not involve patients in other areas of the study design.

### Measures

Our primary outcome, change in daily step counts, will be passively collected by a mobile phone application during the time that patients remain in the intervention. For secondary outcomes, we will derive HbA1c, the average plasma glucose over the previous eight to 12 weeks, recommended as a means to diagnose diabetes(27), from patients’ electronic health records (EHR). We will use the most recent, available measurement from a maximum of 12 months before participating in the study. After 6 months, we will again assess the most recent HbA1c (pulling from patients’ EHR), ensuring that at least 3 months elapsed between baseline and follow-up HbA1c levels. For additional secondary outcomes, we will administer a survey at baseline and 6-month follow-up.

The project coordinator and research assistants will be responsible for managing patient data collection. Data will be stored on UCSF Qualtrics and the HealthySMS platform. Once data from all patients is collected, it will be stored on UCSF’s Secure Box, a secure cloud-hosted platform. See **Table 1** for all included questionnaires.

**Table 1.**

| Measures | Baseline | 6 months follow-up | Weekly |
| --- | --- | --- | --- |
| PHQ-8 | x | x |  |
| RM 1-FM | x | x |  |
| Mood-rating (scale 1 to 9) |  |  | x |
| SEMCD | x | x |  |
| GAD-7 | x | x |  |
| BIS/BAS | x |  |  |
| CAHPS | x | x |  |
| BADS-SF | x | x |  |
| LEIDS-R | x | x |  |
| Neighborhood Scale* | x |  |  |
| UCLA Loneliness 3-item | x |  |  |
| Social Support Scale* | x | x |  |
| Mobile Phone Affinity Scale* | x |  |  |
| IPAQ* | x | x |  |
| System Usability Scale |  | x |  |

The measures above were taken from validated questionnaires. \*The IPAQ: International Physical Activity Questionnaire, RM 1- FM: Physical Activity Stages of Change - Questionnaire, Self-Efficacy for Managing Chronic Disease and System Usability Scale Social Support Scale, LEIDS-R, BADS-SF, and Social Support Scale were modified to match the sociodemographic characteristics of our patient population (i.e rephrasing to decrease literacy levels). The PHQ8, GAD7, BIS/BAS, Mobile Phone Affinity Scale, Neighborhood Scale, UCLA Loneliness 3-item scale, CAHPS scale and System Usability were not modified. We translated the RM 1- FM: Physical Activity Stages of Change - Questionnaire, the Neighborhood Scale, UCLA Loneliness 3-item subscale, and Mobile Phone Affinity Scale questionnaires into Spanish. For the PHQ-8, GAD-7, BADS-SF, Behavioral Avoidance/inhibition (BAS/BIS) scales, Self-Efficacy for Managing Chronic Disease, IPAQ: International Physical Activity Questionnaire, LEIDS-R, CAHPS scale and System Usability Scale we utilized validated translated versions used in previous studies. The Spanish translations of the BADS-SF, LEIDS-R, Self-Efficacy for Managing Chronic Disease and IPAQ: International Physical Activity Questionnaire were modified to match the sociodemographic characteristics of our patient population (i.e rephrasing to decrease literacy levels).

#### Engagement measures

In addition to physical activity and the measures mentioned above, we will also examine engagement measures, such as (1) times that the app was opened, (2) time spent reading the messages, (3) Usability data, assessed by the Systems Usability Scale(28) and open ended qualitative questions about their opinions of the app.

### Procedure

#### Determining content of motivational message categories

We designed motivational text-messages according to a behavioral theory framework, using the Capability, Opportunity, Motivation, Behavior (COM-B) model(29). COM-B is a behavioral change model that proposes that engaging in a particular behavior depends on the dimensions of capability (physical and psychological), opportunity (social and physical) and motivation (need to engage in the behavior more than in other behaviors). Messages were designed to fit into these three dimensions. Before the start of the RCT, English and Spanish speaking patients tested the messages in the Amazon Mechanical Turk (MTurk) crowdsourcing platform. Patients on MTurk were given random messages and asked to categorize them into our overarching theoretical categories. MTurk workers and each coauthor also rated messages for their overall quality from 1 to 5.

#### User Centered Design Testing

We utilized user-centered design (UCD) methods(30) to iteratively develop the content and text messaging system. We conducted three iterative phases of UCD with ten patients each (total n=30). The first phase consisted of 1.5-hour individual semi-structured interviews that took place at the offices at ZSFG. Findings from phase 1 were used to inform content and information delivery decisions of the final intervention including selecting the thematic message categories and the design decisions. The second phase ran as a usability test in which patients tested out an early prototype of the mobile application. The third phase tested out the final DIAMANTE intervention including thematic message content and finalized application in order to address any user-related issues prior to launching the randomized control trial (See **Figure 1** for an overview of the different UCD phases and the RCT).

**Figure 1:**
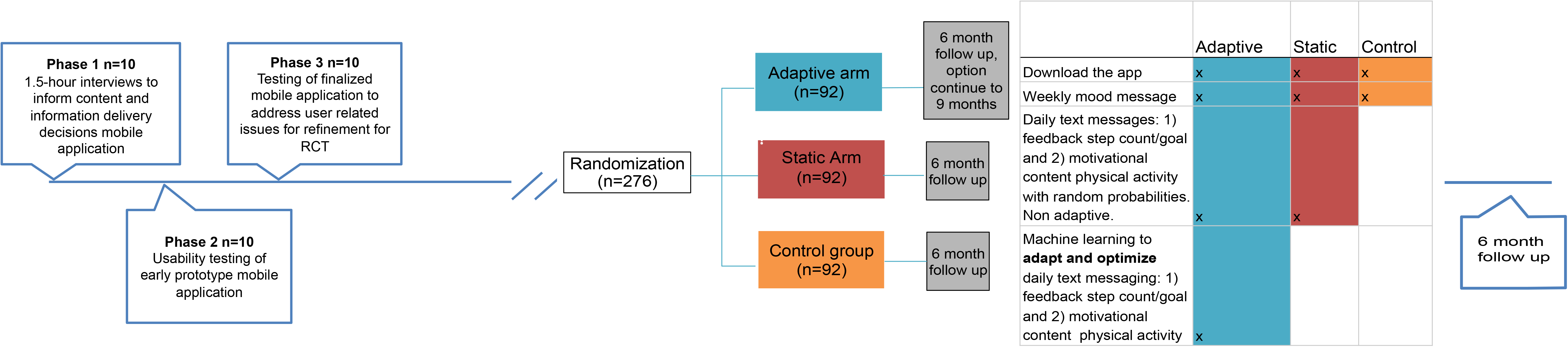
Overview User Centered Design process (UCD) and randomized trial. We first conducted three iterative phases of only UCD with ten patients each (total n=30). The DIAMANTE trial will have different intervention groups: adaptive (n=92), static (n=92) and control (n=92).

#### Interventions

We employ a mobile phone app, “DIAMANTE” developed by Audacious Software https://diamante.healthysms.org/. This application tracks step counts by pooling from Google Fit, Apple HealthKit or the built-in pedometer on patients’ phones. We use a text-messaging platform HealthySMS, developed by Dr. Aguilera, to send text-messages and manage patient responses back to our system. The app only needs to be installed once, but has to remain open consistently. The app is designed in English and Spanish versions and is freely available as a download from the Apple App Store and Android Google Play application.

**Figure 1**. shows the different intervention groups during the trial period. Briefly, both the adaptive and uniform random group will receive the same types of messages: feedback (4 active categories plus no message) and motivation (3 active categories plus no message). However, the message categories, timing and frequency will be optimized by a reinforcement learning algorithm in the adaptive group, and will be delivered with equal probabilities in the uniform random group (following a uniform random distribution). The control group will not receive these messages (only a weekly mood check-in message). All groups will have the app downloaded on their phone and their steps will be passively tracked within the app. See Figure 1 for an overview.

### Control condition

Control patients will only install the app on their phone and will not receive any feedback messages. They will receive one message a week, on a fixed day, asking them to assess their mood in the previous week on a scale of 1 to 9. The message will be sent weekly at 10:00 am. Non-responders will receive reminders to submit their mood self-assessments in two-hour intervals.

### Uniform random message arm

We will send patients up to two messages per day within 4 randomly selected time intervals. These messages are based on the COM-B framework (examples shown in **Table 1 A/B**). In addition, they will receive one message, on the seventh day, which will ask patients to rate their mood on a scale from 1 to 9. Physical activity (step-count/day) will be passively monitored via the app on their smartphone.

### Adaptive message arm

Patients in the adaptive messaging arm will receive the daily COM-B messages (equal to the uniform random arm, examples shown in **Table 1 A/B**), but the message categories, timing and frequency, will not be chosen randomly, but by using a reinforcement learning (RL) algorithm. This allows us to adequately assess whether differences in effects are driven by the use of the RL algorithm.). In addition, they will receive one message, on the seventh day, which will ask patients to rate their mood on a scale from 1 to 9. Physical activity (step-count/day) will be actively monitored via the app on their smartphone.

Patients in all groups will receive reminders to open the app if no data is being transmitted. Additionally, patients in all groups can reply ‘STOP’ or ‘PARAR’ if they wish to stop receiving messages. Finally, the researchers will monitor the incoming step data. The researchers will contact the patients by phone for troubleshooting if patients’ phones are not transmitting data for more than 3 days.

### Adaptive Learning algorithm

We developed a reinforcement learning algorithm (an area of machine learning) based on previous work(31). On a daily basis, this algorithm assesses 1. which feedback message (**Table 2A**) 2. which motivational message (**Table 2B**) and 3. which time period of the day (in intervals of 2.5 hours from 9:00 am to 7:00 pm) is predicted to maximize the number of steps walked the next day. We use algorithms for contextual multi-armed bandit (MAB) problems, as these have been employed in different domains (including mobile health) and show promise to maximize cumulative rewards in sequential decision tasks (here, which sequences of messages optimally promote physical activity)(32). A contextual MAB problem is a reinforcement learning setting in which the algorithm choses between different treatment options which all have different reward functions. The reward functions depend on contextual variables.

**Table 2A.** Different categories with feedback messages that the algorithm chooses from. The algorithm can also choose not to send a message (category no feedback).

| **Feedback messages** | **Examples** |
| --- | --- |
| 0. No Feedback Message |  |
| 1. Reaching goal | *“Yesterday, you did not reach your goal”* |
| 2. Steps walked yesterday | *“Yesterday, you walked 3824 steps”* |
| 3. Walked more/less today than yesterday | *“Yesterday, you walked more than your goal”* |
| 4. Steps walked yesterday, plus a positive/negative motivational message | *“You walked 8000 steps yesterday. Great job!”* |

**Table 2B.**
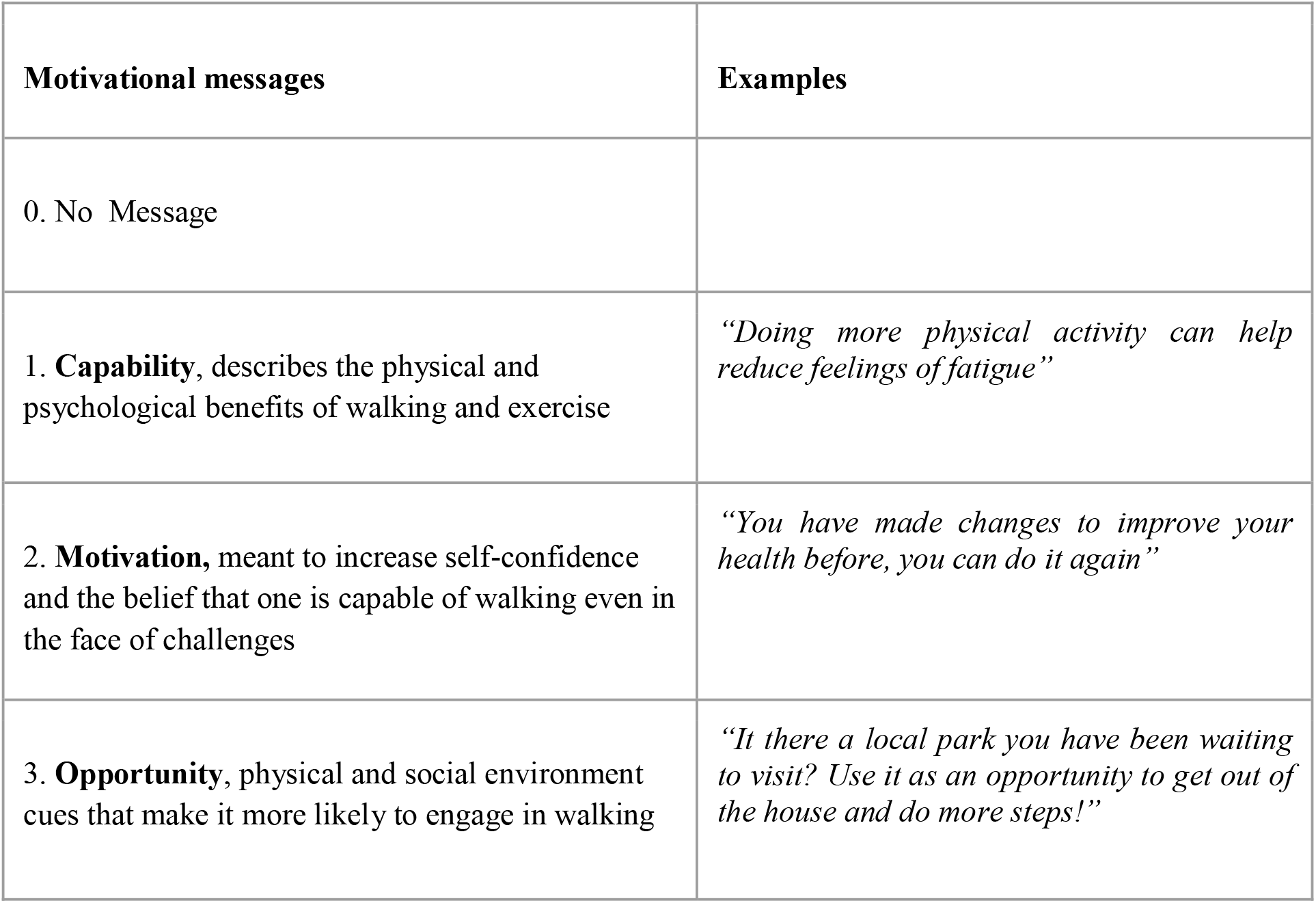
Different categories with Motivational messages that the algorithm chooses from. The algorithm can also choose not to send a message (category no feedback).

We use Thompson Sampling, a Bayesian method, which will allow us to continuously learn which feedback and motivational messages are effective for a user, based on contextual features like their previous physical activity, demographic and clinical characteristics (such as age, gender and PHQ-8 scores). Thompson sampling can effectively deal with small amounts of data and addresses the exploration/exploitation trade-off(33). As such, it frequently picks out from the most rewarding messages and occasionally explores the messages with uncertainty in their reward.

More specifically, each morning, the algorithm evaluates which messages will likely increase the physical activity for every participant in the upcoming day, and at which time period the messages should be delivered. The algorithm training data consists of the historical data of all participants (contextual variables), which include which messages were sent previously and within which time periods, and select clinical/demographic data (such as age, language and depression scores) to improve prediction abilities.

During the trial, patients will receive messages for 6 months. After 6 months, we will provide patients the possibility to remain receiving messages for up to 9 months.

### Statistical analysis plan

#### Baseline characteristics

We will provide descriptive summaries and examine any potential imbalances between the intervention and control groups using χ^2^-tests for categorical variables and independent samples t-tests for continuous variables.

#### Primary and secondary hypothesis

Analyses will be conducted on an intention-to-treat basis. We will use full-information maximum likelihood to handle missing data, which has been shown to be preferred over multiple imputations(34). We will include patients in the analysis for primary and secondary outcomes that have at least one month of data available. The primary outcome measure, increase in daily step counts, and the secondary outcomes measures (HbA1c, PHQ-8 and other questionnaire data, see measures) will be analyzed using a conditional growth model, in line with previous work(35), with intervention (adaptive, uniform random, control) being the fixed effect and time being nested within individuals. Let *j = 1*, …, *n, i = 1*, …, *t*, where *t* is the number of recorded days for the *j*th patient Y_ij_ is the daily steps count on the *i*th day of *j*th patient.

Let A_j_ = 2, 1 or 0 for patient *j* in adaptive, uniform random or control arm, and let T_ij_ be the *i*th day for patient *j*.

The daily steps for patients given the treatment, in equation form:

At level 1 (within-patient):

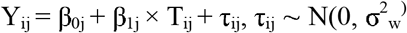

At level 2 (between-patient):

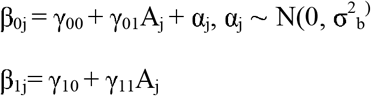

As this is a randomized trial, we will not adjust for any patient characteristics or patient engagement measures in the primary intention-to-treat analysis unless there is a significant imbalance between the arms of the trial.

### Pre-planned subgroup analyses

We will conduct an exploratory sub-group analyses for our primary and secondary hypotheses for English versus Spanish speaking patients. We expect at least 50% of the sample to be Spanish speakers given our previous testing recruitment.

#### Power Calculations and Sample Size

Power for growth models were calculated based on Monte Carlo simulations using the ‘*nlme*’ package in R(36) We conducted simulations, using data from our own study team testing the app (n=47) and data from pilot patients (n=8) to estimate the model parameters for calculating the necessary sample size. We used an estimated mean increase of 1250 steps based on previous work (37, 38). With 5000 Monte Carlo simulation runs, number of days for each patient *t* ∼ *Uniform(28,180)*, we estimate that we need n = 160 for 2-arms (the adaptive vs. uniform random) for 80% power. Inflating to include the control arm and controlling for 15% drop-out, we aim to recruit a total of 276 patients. We believe that low drop-out is feasible as (1) the primary outcome of interest will be passively assessed and thus requires limited proactive engagement by the user, and (2) we have a comprehensive protocol to automatically text users who have closed the app or are not transmitting data.

#### Compensation

Patients will receive a compensation of $40 for participation in the baseline part of the study, and an additional $70 for completion of the 6-month follow up.

#### Ethics and Dissemination

The UCSF Institutional Review Board has approved this protocol. All protocol amendments will be communicated for approval to the USCF IRB. We will ensure that our text messaging content is publicly available through a creative commons licensing agreement. The HealthySMS system is available for use upon request.

#### Data statement

We will submit study-results for publication in peer reviewed journals and presentation at (inter)national meetings, taking into account relevant reporting guidelines (e.g. CONSORT(39)). We will attempt to publish all findings in open-access journals when possible, or in other journals with a concurrent uploading of the manuscript content into PubMed Central for public access.

Curated technical appendices, statistical code, and anonymized data will become freely available from the corresponding authors upon request.

### Safety protocol

We will review the clinician dashboard for patients in all arms of the trial to identify safety concerns that may emerge over the course of the trial. In addition, we will be automatically notified when key events, such as self-reported suicidality (messages with the words “Kill, Die, Suicide, Don’t want to live, Do not want, Bridge, What is the point, Harm, Hurt, Gun” for English speaking patients; and “Tiro, Matar, Morir, Suicidio, suicidar, no quiero, puente, para que, daño, dañar, dano, danar” for Spanish speaking patients) occur. We will provide immediate phone outreach and will refer patients to urgent or emergency care as necessary. Dr. Aguilera will be available to consult and intervene as necessary for mental health emergencies.

## DISCUSSION

In this randomized controlled trial, we aim to examine the effect of a smartphone app that uses reinforcement learning to predict the most effective messages for increasing physical activity in 276 low-income, ethnic and racial minority patients with diabetes and depression in urban public sector primary care clinics. We will compare this intervention to uniform random messages, delivered with equal and unchanging probabilities, and a control group that only receives a weekly mood message.

### Decreasing health disparities

Though the numbers of mHealth pilot studies are increasing in vulnerable populations, many of these fail to follow through with an implementation component to the study design(40). Here, we are using a blended design: while the intervention is in addition to current care, there are ways we are attempting to make it more a part of patients’ clinical care. For instance, patients are mainly approached through primary care health providers, which recommend eligible patients whom they think are directly interested in a physical activity intervention. In addition, we will make patients’ data available to providers: a summary of the step increase for that patient at the conclusion of the study and updated PHQ-8 and GAD scores entered into the record. Our study therefore is the first step to addressing this gap because of its integration in primary care clinics that serve low-income patients. Future work should focus more specifically on implementation of the app as part of routine clinical care.

### Physical Activity measure

We chose passively collected daily step count from patients’ pre-owned digital devices as a measure of physical activity. Although there are many different ways to measure physical activity, daily step count seems to be a particularly relevant measure because of: 1. Its relative ease to measure, 2. The clinical importance of individuals’ walking behavior. Low number of daily step counts have been associated with all-cause mortality in some longitudinal studies(41) and results from pooled population studies show clear dose–response effects of physical activity to overall mortality(42). In patients with Diabetes Type 2, several studies have now shown that increasing step counts can significantly decrease HbA1c levels. For instance, a 10,000 steps per day walking prescription increased steps and decreased HbA1c in patients with Diabetes Type 2(43). Further, Manjoo et al. found that each standard deviation increase in daily steps was associated with a 0.21% decrease in HbA1c(44). However, negative findings have also been reported. For instance, a meta-analysis by Qui et al. found that step-counter use was associated with increased steps per day (over 1800 more steps compared to a control group) among people with diabetes, but not with lowering of HbA1c(45).

In exploratory post hoc analyses, we will also be able to examine the more immediate effect of physical activity messages, e.g. on hourly steps in addition to daily steps, which will help to improve future physical activity interventions (e.g. deliver messages at the right times). For instance, it is possible that one could receive a message in the morning and make plans to walk in the afternoon or evening, or messages could have more of an immediate impact. This information is currently unknown.

### Personalization of intervention

The results of this RCT will help us understand if adaptive mHealth interventions for depression and diabetes are more beneficial than interventions that do not use learning algorithms. If mHealth interventions are not personalized, their efficacy might be low, due to low engagement and high drop-out rates(20). The use of machine learning to adapt interventions according to users’ characteristics and behaviors is still in its early stages, but shows promise(46). For instance, Yov-Tom et al. using an adaptive learning algorithm, found that adaptive feedback messages were more effective in increasing the amount and speed of physical activity and also reduced HbA1c in sedentary patients with diabetes type 2(31). Further, Zhou et al. showed short-term efficacy of using adaptive weekly step goals determined by reinforcement learning in healthy patients(47). The current study, with a relatively large group of patients, will further increase our understanding of the potential of machine-learning driven text messaging interventions.

## Limitations and strengths

### Limitations

First, the results of this study might be specific to this population of low-income ethnic minority patients and might therefore not be generalizable to other populations. However, the inclusion of vulnerable patients in a primary care setting increases the likelihood that this intervention will be effective for other underserved populations. Further, our study procedures do not allow a double-blind design, as researchers and patients need to be made aware of the nature and frequency of messages they are receiving. Additionally, as with all digital interventions, technical issues might arise leading to unreliable step-count data and reduced ability for the algorithm to predict the most effective messages. The researchers and technical personnel will frequently check our server for data collection troubleshooting. Patients will also be made aware when their data is not collected correctly.

### Strengths

Diabetes and depression are among the top ten causes of disability in the US(48). Developing cost-effective and scalable models of care for patients with common chronic conditions has been postulated as of key importance in improving the performance of health care systems(49). If mHealth apps that target diabetes and depression through their common risk-factor physical inactivity are effective, they can have a major public health impact. Further, because the learning algorithm that we apply in this study is automated and delivers adaptive messages based on patients’ behaviors, it can potentially be applied in other patient populations with a wide range of conditions.

## Conclusion

The outcome of this trial will provide information on the effectiveness of a text-message based smartphone app that uses machine learning to increase physical activity in low-income ethnic minority patients in primary care settings. The results will provide key information on the effectiveness of adaptive mobile applications, compared to more traditional static digital interventions. If effective, this application has the ability to decrease healthcare disparities by providing a type of personalized care to a diverse and traditionally hard to reach group of underserved patients.

## Author contributions

Dr. Aguilera and Dr. Lyles designed the overall study. Ms. Hernandez-Ramos, Ms. Gomez-Pathak, Mr. Miramontes, Ms. Cemballi, Dr. Sarkar and Dr. Figueroa were involved in the design and implementation of the User Centered Design phase and RCT. Dr. Jay Williams, Ms. Modiri, Mr. Aggarwal, Dr. Figueroa and Dr. Yom-Tov were involved in the design of the reinforcement learning algorithm. Dr. Figueroa wrote the first draft of the paper. Dr. Chakraborty, Ms. Yan and Dr. Xu revised the statistical analyses and power calculation sections of the paper. All authors revised the manuscript for relevant scientific content and approved the final version of the manuscript.

## Acknowledgements

We would like to acknowledge Chris Karr who helped develop and manage the automated text messaging service, passive data collection and reinforcement algorithm and Patricia Avila Garcia who was involved in the design and implementation of the User Centered Design Phase of the study.

